# Leveraging human genetic variation to therapeutically target hundreds of genes with dominant & dispensable disease alleles

**DOI:** 10.64898/2026.03.26.26349431

**Authors:** Grace D. Ramey, Quinn T. Cowan, Akshita G. Saxena, Bria L. Macklin, Hannah L. Watry, Xiaoyue Mei, Philip Dierks, Luke M. Judge, Bruce R. Conklin, John A. Capra

**Author notes:** Co-Correspondence.

## Abstract

Here we identify a novel therapeutic opportunity for over 500 genes with putative “dominant & dispensable” (D&D) disease alleles. In these haplo*sufficient* genes, a single functional allele may be sufficient for health, presenting the opportunity for therapeutic approaches that silence the pathogenic allele. We show that allele-specific targeting of common heterozygous genetic variation linked to D&D alleles enables a disease mutation-agnostic gene therapy approach that increases the number of patients treatable with a single therapy. In some disease genes, this approach would allow >80 times as many patients to be treated as mutation-specific strategies. D&D alleles cause diverse diseases, including neurodegeneration, cardiomyopathies, retinopathies, and diabetes, demonstrating the therapeutic opportunity of this approach across physiological systems. To enable broad application of allele-specific mutation-agnostic targeting, we provide genome-wide maps of common heterozygous variants that support D&D disease allele disruption by multiple CRISPR-based editing technologies, including Cas9 nucleases, base editors, and epigenome editors.

## Introduction

Dominant negative and gain-of-function (DN/GOF) mutations cause hundreds of diseases that affect diverse systems across the body^1^. In these conditions, a mutated gene copy interferes with or overrides the function of the wild-type allele (Figure 1A). DN/GOF diseases are particularly challenging to treat by gene replacement and other pharmacological approaches because promoting the function of the healthy allele often does not suffice to prevent the toxic effect of the DN/GOF allele^2^.

**Figure 1.**
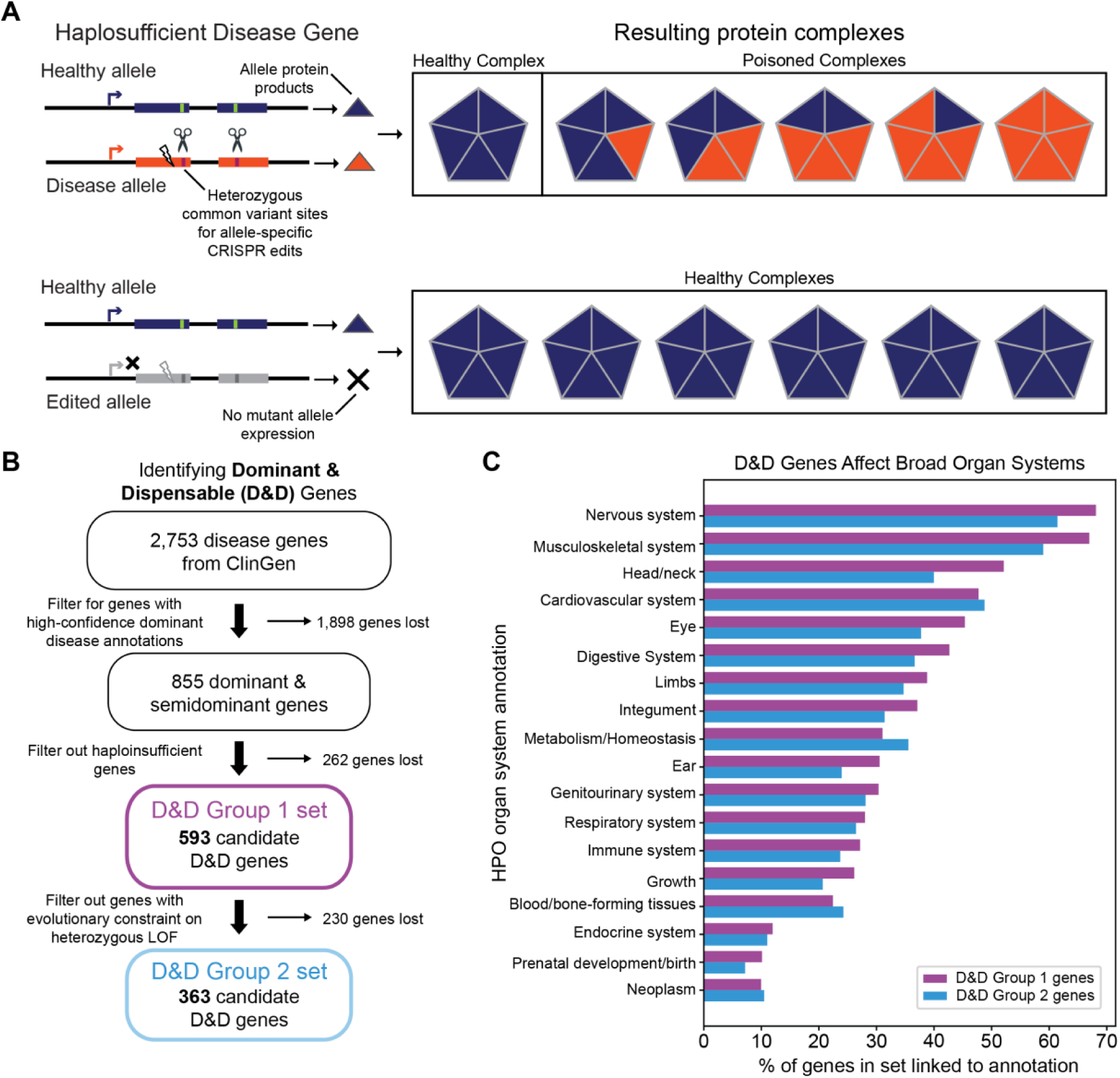
Dominant and dispensable (D&D) disease alleles are candidates for allele-specific therapeutic gene editing. (A) Schematic of a dominant negative (DN) disease allele in a haplosufficient disease gene. Lightning bolt represents a pathogenic disease mutation. The scissors represent potential allele-specific edits targeting common heterozygous variation. Triangles represent protein products that form a protein complex. The complex is nonfunctional, or ‘poisoned’, when a protein produced from the disease allele is included. Because the gene is haplosufficient, selectively disrupting the disease allele can restore a healthy phenotype. We refer to such alleles as dominant & dispensable (D&D). (B) Genes with D&D alleles were identified using ClinGen annotations and evidence of selection against heterozygous loss-of-function variants (shet). (C) Dominant conditions associated with D&D genes span a broad range of phenotypes and physiological systems, with nervous system abnormalities as the most prevalent. Annotations come from the Human Phenotype Ontology (HPO). LOF = loss-of-function.

The potential for precision genome editing provided by CRISPR technologies has enormous promise for treating diseases caused by DN/GOF mutations^3,4^, as targeting sequences that differ by a single nucleotide can enable allele-specific disruption of the DN/GOF disease allele^5–7^. However, clinical application of this approach faces significant obstacles. In particular, many diseases result from hundreds of different DN/GOF mutations in the same gene, so it is prohibitively costly to pursue clinical trials and FDA approval for mutation-specific therapies, many of which only treat a handful of patients each^8–10^. Therefore, disease mutation-agnostic approaches are needed to make CRISPR gene therapies financially feasible and applicable to broad patient populations.

We have identified a therapeutic opportunity to address this challenge and treat hundreds of DN/GOF diseases at population scale by leveraging patterns of common genetic variation in humans. Our approach selectively targets the allele carrying the DN/GOF mutation for inactivation using common variants in or near the gene body, leaving the healthy allele intact (Figure 1A). For this approach to work, the gene must be haplosufficient, i.e., the remaining functional allele must be able to restore a healthy phenotype on its own. In other words, the gene copy with the mutation must be *dispensable* due to the ability of the remaining allele to sufficiently compensate.

We refer to disease genes targetable by this approach as “dominant & dispensable” (D&D) to reflect the fact that their <u>d</u>ominant disease alleles (DN or GOF) are targetable and ultimately <u>d</u>ispensable due to the gene’s haplosufficiency. Thus, they can be inactivated for therapeutic purposes. This precise allele-specific genome editing strategy has already begun to show promise for individual diseases and genes, such as Charcot-Marie-Tooth disease caused by mutations in *NEFL*^11–13^.

Here we show that allele-specific editing of D&D alleles has potential to be widely applied to diverse conditions caused by DN/GOF mutations. Integrating clinical, genomic, and evolutionary methods, we found that targeting common variation to inactivate DN/GOF disease alleles is a highly scalable approach applicable to hundreds of disease genes. Genes with D&D alleles are associated with a wide variety of conditions, including diseases of the central nervous system, heart, lung, and many other tissues. To enable our approach, we identified common heterozygous sequence variants that support mutation-agnostic, allele-specific inactivation of the DN/GOF disease alleles at scale in the human population. Our findings demonstrate the feasibility of allele-specific editing and provide a road map for the development of treatments for large numbers of patients with debilitating genetic conditions that currently have no effective therapies.

## Results

### Hundreds of disease genes are D&D and thus candidates for therapeutic allele-specific inactivation

For a disease allele to be D&D, the affected gene must be haplosufficient in the cell types where allele-specific inactivation is needed. Haplosufficiency is difficult to define as it exists on a functional spectrum that can be context dependent. The standard definition of haplosufficiency is the “condition where having one functional and one nonfunctional version of a gene provides sufficient gene activity to give a normal phenotype”^14^. However, “normal” can be ambiguous in human health and physiology. In an ideal case, the loss of one allele would have no measurable effect. Yet the loss of a single allele will often have a small effect that is still in the normal physiological range and could still be considered haplosufficient. For the purposes of our study, we define “normal” broadly, with the assumption that the consequences of allele inactivation on gene dosage would be balanced against the potential benefit of inactivating the disease allele. Like many therapies, this will have to be evaluated for each disease.

Given the challenges in defining haplosufficiency and the spectrum on which it exists, we integrated large clinical genomic databases and genetic variation from population-scale genome sequencing cohorts to identify genes with candidate D&D disease alleles genome-wide. Databases such as ClinGen and OMIM provide reports of disease phenotypes, mode of disease inheritance, and dosage sensitivity for many human disease genes. The advent of large databases of whole-genome sequences from diverse individuals, such as gnomAD, also enables estimation of evolutionary constraint against loss-of-function variants. These clinical and evolutionary data capture different aspects of haplosufficiency useful for identifying D&D candidates. Using ClinGen data, we focused on genes with a high-confidence association with dominant or semidominant disease inheritance and limited evidence of dosage sensitivity. Genes are annotated as “dosage sensitive” if a loss or inactivation of one allele (e.g., via a loss-of-function mutation) is known to cause disease. Thus, to identify putative genes with D&D alleles, we considered genes lacking any “dosage sensitive” annotation, given that one allele must be sufficient for a functional phenotype.

Out of ∼2,700 disease genes (covering ∼1,900 conditions) in the ClinGen database, we identified 593 gene candidates with D&D disease alleles (Figure 1B). For brevity, we use the term “D&D genes” for genes with D&D disease alleles, but we note that dominant describes the disease-causing alleles and their associated phenotypes rather than the genes themselves. We refer to this set of 593 putative candidates as D&D Group 1 genes, and unless otherwise specified, “D&D genes” refers to this group.

Since dosage sensitivity annotations in ClinGen are still in development and incomplete, we also incorporated an evolutionary metric of <u>s</u>election against <u>het</u>erozygous loss-of-function variants (shet). We reasoned that genes that can tolerate heterozygous loss-of-function mutations are more likely to be haplosufficient in the context of human evolution^15,16^. Filtering D&D Group 1 genes to those with less than a 10% reduction in fitness upon heterozygous loss of function (shet < 0.1) yielded 363 genes (Figure 1B). However, we note that this approach likely excludes some genes that are haplosufficient in the cell types where disease manifests. We refer to genes meeting both the ClinGen and evolutionary criteria as D&D Group 2 genes (Figure 1B). Given the complexity of defining haplosufficiency in clinical contexts, the D&D gene sets are likely to change as disease-gene annotations improve and the understanding of haplosufficiency for genes in specific cell types becomes more apparent.

Many genes that cause debilitating diseases are present within the D&D gene sets (Table 1). For example, genes linked to nervous system conditions such as channelopathies (*SCN1B*, *KCNJ2, KCNJ5*) and peripheral nerve degenerative diseases such as Charcot-Marie-Tooth disease (*MFN2, NEFL*) have D&D alleles. Genes for serious musculoskeletal and cardiovascular conditions, such as achondroplasia (*FGFR3*) and hypertrophic cardiomyopathy (*MYH7*), are also present. D&D mutations are also found in many immune genes, such as *STAT1*, that cause immunodeficiencies. Many of these diseases, e.g., motor neuron diseases, are in need of therapies, including Charcot-Marie-Tooth disease, which is linked to 15 D&D genes.

**Table 1.**
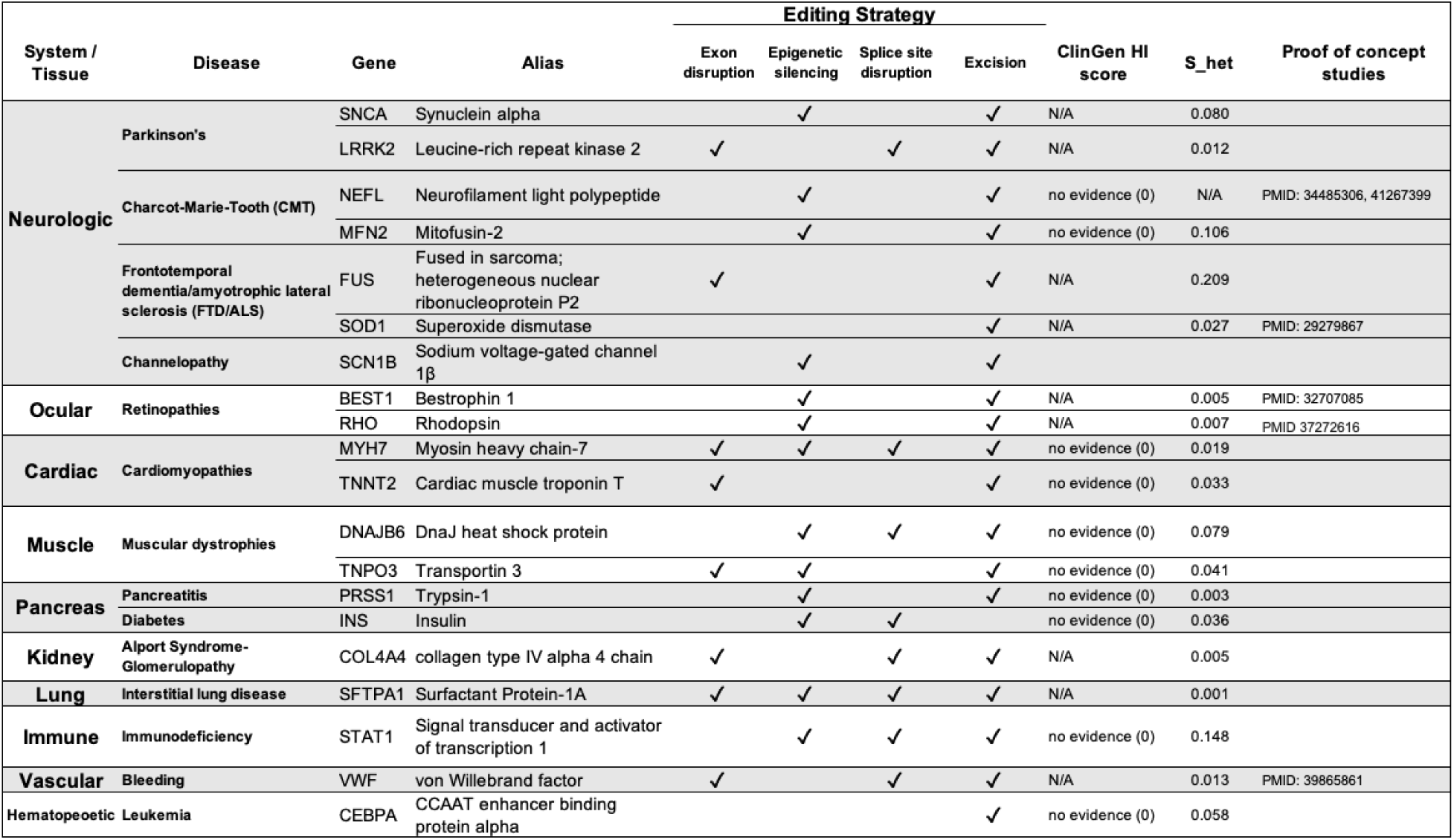
Dominant and dispensable (D&D) disease alleles represent an untapped therapeutic opportunity. Examples of genes with D&D disease alleles that could benefit from allele-specific disruption of pathogenic mutations. D&D disease alleles cause disease across diverse organs and tissues, with neurological diseases being the most common (Figure 1C). Multiple genes with D&D alleles are linked to conditions such as Parkinson’s disease, Charcot-Marie-Tooth disease, and frontotemporal dementia (FTD)/amyotrophic lateral sclerosis (ALS). N/A = not annotated in ClinGen Dosage Sensitivity Curation or shet list.

### D&D alleles cause disease in diverse organs and tissues

Dominant diseases linked to the 593 D&D Group 1 genes affect diverse physiological systems based on annotations from the Human Phenotype Ontology (HPO) catalog (Figure 1C). Nervous system symptoms were most common among linked dominant diseases; 404 of the D&D Group 1 genes (68%) are annotated with “abnormality of the nervous system”. Abnormalities of the musculoskeletal system, head/neck, cardiovascular system, and the eye are also each associated with more than 45% of the D&D Group 1 genes. These percentages sum to greater than 100% because diseases often affect multiple systems. We observed a similar distribution across physiological systems for the D&D Group 2 genes (Figure 1C). While the distribution of D&D allele effects across bodily systems is likely influenced by biases in our knowledge of the genetic basis and manifestation of diseases, these results underscore the breadth of the therapeutic opportunity for D&D alleles.

To address overrepresentation biases, we tested whether any physiological systems or functional annotations were enriched among D&D genes compared to all disease genes.

Among physiological systems annotated by HPO, diseases affecting the nervous system, cardiovascular system, and musculoskeletal system were enriched by > 15% (FDR-corrected p < 0.001, Supplementary Table 1). Additionally, diseases of the blood/bone-forming tissues, eye and ear, and immune system were all enriched by 8% or more (FDR-corrected p < 0.001, Supplementary Table 1) in the D&D set. In the D&D Group 2 set, abnormal cardiovascular, nervous system, musculoskeletal, and blood/bone phenotypes were also enriched each by over 9% (FDR-corrected p < 0.05, Supplementary Table 1).

Many molecular function and biological process annotations from the Gene Ontology knowledgebase were also enriched among the D&D genes. For example, among molecular function terms, protein homodimerization activity (GO:0042803, FDR-corrected p < 0.001), ion channel activity (GO:0022843, FDR-corrected p<0.001), transcription factor binding (GO:0140297, FDR-corrected p < 0.001), and GTPase activity (GO:0005525, FDR-corrected p < 0.001) were all strongly enriched (Supplementary Figure 1A). The enrichment for dimerization, transcription factors, and ion channels is consistent with known mechanisms for dominant negative alleles^1^. Additionally, supporting the cardiovascular and musculoskeletal HPO enrichments, many cardiac and muscle GO processes were enriched in the D&D gene set, including heart contraction (GO:0060047, FDR-corrected p < 0.001) and ventricular cardiac muscle tissue morphogenesis (GO:0055010, FDR-corrected p < 0.001) (Supplementary Figure 1B). Full enrichment results are reported in Supplementary Figure 1 and Supplementary Data Table 1.

### Genes with D&D alleles harbor many distinct pathogenic mutations

If only one or a small number of DN/GOF mutations are observed in a gene, then directly targeting these alleles for gene editing with tailored CRISPR reagents is the most straightforward therapeutic approach. In contrast, if a gene has many DN/GOF mutations, then it is impractical to develop a large number of targeted therapies for each mutation one-by-one. Given the large potential for therapeutic silencing of disease-causing mutations in hundreds of D&D genes, we sought to estimate how many mutation-specific therapies would be required for each D&D gene based on its observed patterns of pathogenic mutations.

Analyzing pathogenic mutation annotations in ClinVar for the D&D gene sets, we found that D&D Group 1 genes harbored nearly 27 dominant pathogenic mutations on average, and more than 75% of genes harbored four or more pathogenic mutations (Figure 2A). D&D Group 2 genes harbored a similarly high distribution of mutations per gene (Supplementary Fig 2). As clinical sequencing and our knowledge of disease pathology increases, the number of disease mutations is likely to increase. Indeed, some well-studied D&D genes, such as *MYH7,* have over 140 annotated dominant pathogenic mutations^17,18^. In *MYH7*, these mutations occur throughout the gene in different functional domains. These results illustrate the challenge of mutation-specific gene editing: hundreds of different gene editing treatments could be necessary even for a single gene, and, thus, mutation-independent strategies for targeting D&D alleles are needed (Figure 2B,C).

**Figure 2.**
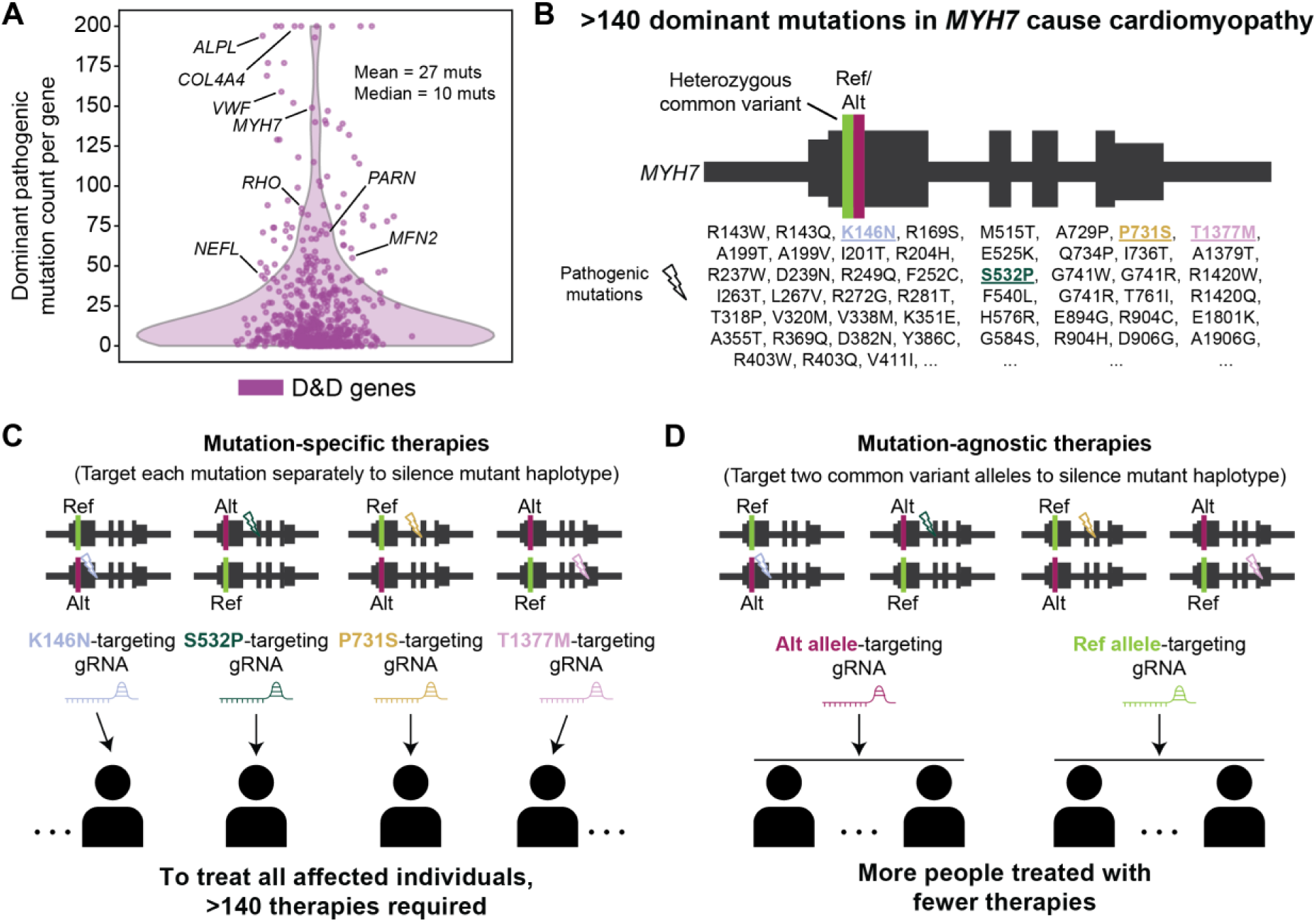
Disease mutation-agnostic editing opportunities enable improved treatment capacity. (A) Dominant pathogenic mutation counts per D&D gene. Most D&D genes have 4 or more mutations, and many have upwards of 100, making mutation-specific therapies infeasible. Counts were truncated at 200 mutations. (B) Example D&D gene *MYH7* in which over 140 different dominant mutations are known to cause cardiomyopathy. Due to the large number of mutations, *MYH7* is a good candidate for mutation-agnostic editing, as described in (C) and (D). It also exhibits a common variant with reference (Ref, green) and alternate (Alt, maroon) alleles in its exons. (C) Schematic of disease-mutation-specific editing applied to *MYH7*. Lightning bolts represent distinct pathogenic mutations. A different gRNA, or therapy, is required to target and therapeutically edit each pathogenic mutation. Therefore, over 140 therapies would be required to treat all affected individuals, which is infeasible. (D) Schematic of disease mutation-agnostic editing for *MYH7*. Even though distinct pathogenic mutations are still present, they fall on the same haplotype as either the reference allele or alternate allele of a heterozygous common variant. Targeting these alleles with gRNAs to silence the mutant haplotype results in many more patients treated with fewer therapies. Note that *MYH7* is not drawn to scale or accuracy here.

### Common genetic variation provides an opportunity for scalable editing in over 95% of D&D genes

The human population carries tens of millions of common, usually benign, genetic variants that are present at high frequencies (>1%) throughout the population. Thus, individual human genomes are heterozygous at ∼1.5-3 million sites^19,20^. These common heterozygous variants provide targets that could enable allele-specific disruption of any nearby D&D alleles without the need to target a specific pathogenic mutation. The ability to therapeutically target D&D alleles with gene editing approaches based on common variation could enable large proportions of individuals to be treated with relatively few therapies (Figure 2D).

Allele-specific gene therapies, such as CRISPR, antisense oligonucleotides (ASOs), and small interfering RNAs (siRNAs) have shown great potential to disrupt disease alleles^11,21–23^. Because CRISPR gene editing provides the greatest allele specificity^24,25^, we focus on the ability of CRISPR methods to target common variants and disrupt D&D alleles, but our strategy is applicable to any allele-specific editing technology. We considered four CRISPR-based allele-specific gene inactivation methods (Figure 3A). The first strategy, *exon disruption*, uses a guide RNA (gRNA) that specifically targets one allele of a heterozygous variant in the coding region of a gene. An insertion or deletion is introduced at the cut site that shifts the reading frame of the allele to induce nonsense-mediated decay (NMD). The second strategy, *gene excision*, produces an allele-specific inactivating excision using a pair of gRNAs targeting heterozygous variants that flank critical functional regions of the gene^11^. For this strategy, we considered pairs of flanking variants within 50 kb of the gene that would not disrupt exons of other neighboring genes. The third strategy, *splice site disruption*, uses a base editor targeted to a heterozygous variant near a splice site to disrupt the splice donor/acceptor sequence on a specific allele. This induces NMD via frameshifts generated by intron retention or aberrant splicing^26,27^. For this strategy, we considered heterozygous variants located roughly within 3-20 bp from a splice site for optimal base editing. The fourth strategy, *epigenetic silencing*, induces heritable allele-specific epigenetic silencing by targeting a catalytically dead Cas9 (dCas9) fused with an epigenetic repressor to a heterozygous variant in the promoter of a gene. This strategy requires a heterozygous variant in a GC-rich region within +/- 500 bp of the transcription start site^28,29^. Each of these approaches can achieve selective inactivation of an allele via targeting the common heterozygous variant without the need to target the disease-causing mutation itself. Thus, all have the potential to provide mutation-agnostic editing.

**Figure 3.**
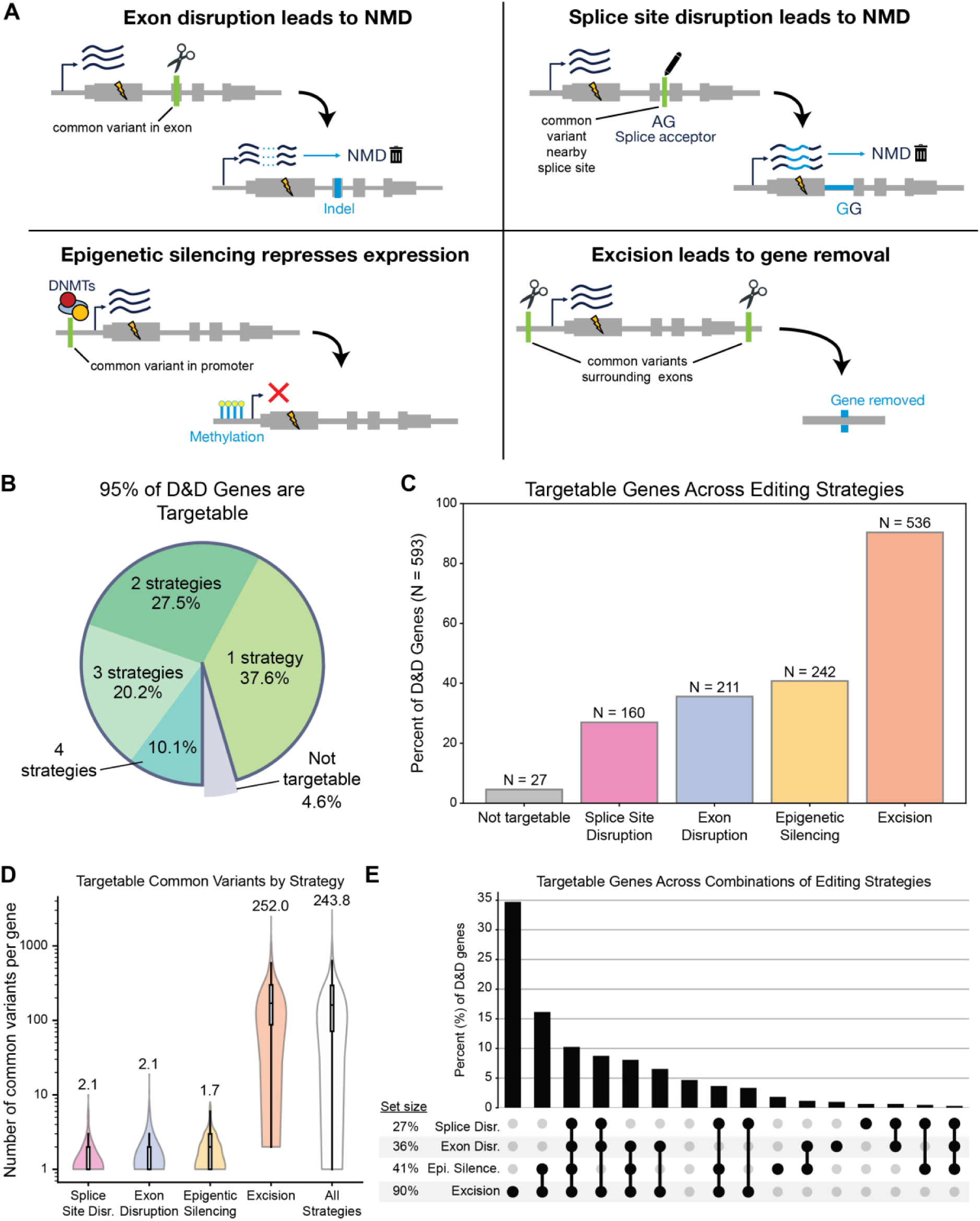
Common-variant-mediated gene editing strategies enable targeting of >95% of D&D genes. (A) We investigated four potential strategies for allele-specific gene editing. Lightning bolts represent disease mutations. Note that the disease mutation must fall in *cis*, i.e., be on the same haplotype, as the targeted variant for the disease allele to be disrupted. The type of targetable heterozygous common variant relevant to each strategy is shown in green, and light blue represents the change that occurs after editing. Each strategy targets a different genomic region (exons, splice sites, promoters, and intergenic regions) resulting in allele-specific disruption or silencing of an allele via multiple outcomes (NMD, epigenetic silencing, or gene removal). (B) Over 95% of D&D genes were targetable with at least one of the four editing strategies, and over half of the D&D genes (54%) were targetable by more than one strategy. (C) Percent of D&D genes targetable by each editing strategy. Based on common variant patterns, excision could target the most genes (90%), followed by promoter methylation (41% of genes), exon disruption (36% of genes), and splice site disruption (27% of genes). (D) Numbers of common variants for each D&D gene targetable by each editing strategy, indicating many opportunities to target each gene. Numbers above violins are means. Boxes represent median and interquartile range (IQR), and whiskers represent 1.5xIQR. (E) D&D genes are often targetable by multiple strategies, providing therapeutic versatility. NMD = nonsense-mediated decay, Disr = Disruption, Epi Silence = Epigenetic silencing.

To quantify the potential for mutation-agnostic, allele-specific inactivation of D&D disease alleles in a broad patient base, we identified very common (global allele frequency ≥ 10%) heterozygous variants in genome-wide sequencing data from the 1000 Genomes Project that could be used to target genes with D&D alleles with the four allele-specific editing approaches. While we focused on variants specific to these approaches, the number of targetable variants and individuals will expand even further as new genome editing methods are developed.

Of the D&D Group 1 genes, 95% (566/593) were targetable by at least one editing strategy (Figure 3B) based on the presence of at least one targetable variant or variant pair linked to the genes. The excision strategy was able to target the largest fraction of D&D genes (Figure 3C), 90% (563/593), due to the greater flexibility in the targeted variants’ genomic locations compared to other strategies (Figure 3A). In particular, we looked within 50 kb windows flanking the gene to identify variants targetable by excision, which is much less restrictive and involves genomic regions under less constraint than, for example, coding regions for the exon disruption strategy. Indeed, more variants were targetable per gene for the excision method than any other strategy (Figure 3D), and the excision method had a mean of 252 variants across excision-targetable genes. In contrast, a mean of around 5 variants per gene were targetable for non-excision strategies (Figure 3D). The next most widely applicable strategy after excisions was epigenetic silencing, which was able to target 41% of genes (242/593) (Figure 3C). Exon disruption in protein coding regions was possible in 36% of D&D genes (211/593), and finally splice site disruption could target 27% of genes (160/593) (Figure 3C). The fractions of targetable genes for each method were similar for the D&D Group 2 gene set (Supplementary Figure 3).

More than half (58%) of D&D genes were targetable by more than one strategy (Figure 3B, E). This flexibility is encouraging because the factors influencing gene editing success are complex and frequently cannot be anticipated in advance of experiments. Therefore, having multiple targets and editing strategies available provides therapeutic versatility.

## Common variants enable therapeutic targeting of D&D genes in over 80% of the population on average

In the previous section, we showed that most D&D genes have common variation that would enable treatment by allele-specific inactivation in heterozygous individuals. To quantify the fraction of *individuals* that could be treatable for each D&D gene, we analyzed common variant patterns in whole genome sequences from the 1000 Genomes cohort. For each of the four editing strategies, we computed how many individuals were heterozygous at each targetable common variant (or common variant pair) and took the union to determine the total number of unique individuals treatable across all editing strategies. In other words, we investigated how many people were heterozygous for at least one common variant targetable by at least one editing strategy per gene (Figure 4A). This approach assumes that rare disease alleles occur randomly across haplotype backgrounds. While this will not always be true (e.g., in founder effects), it provides a proxy for the ability to edit alleles on diverse backgrounds.

**Figure 4.**
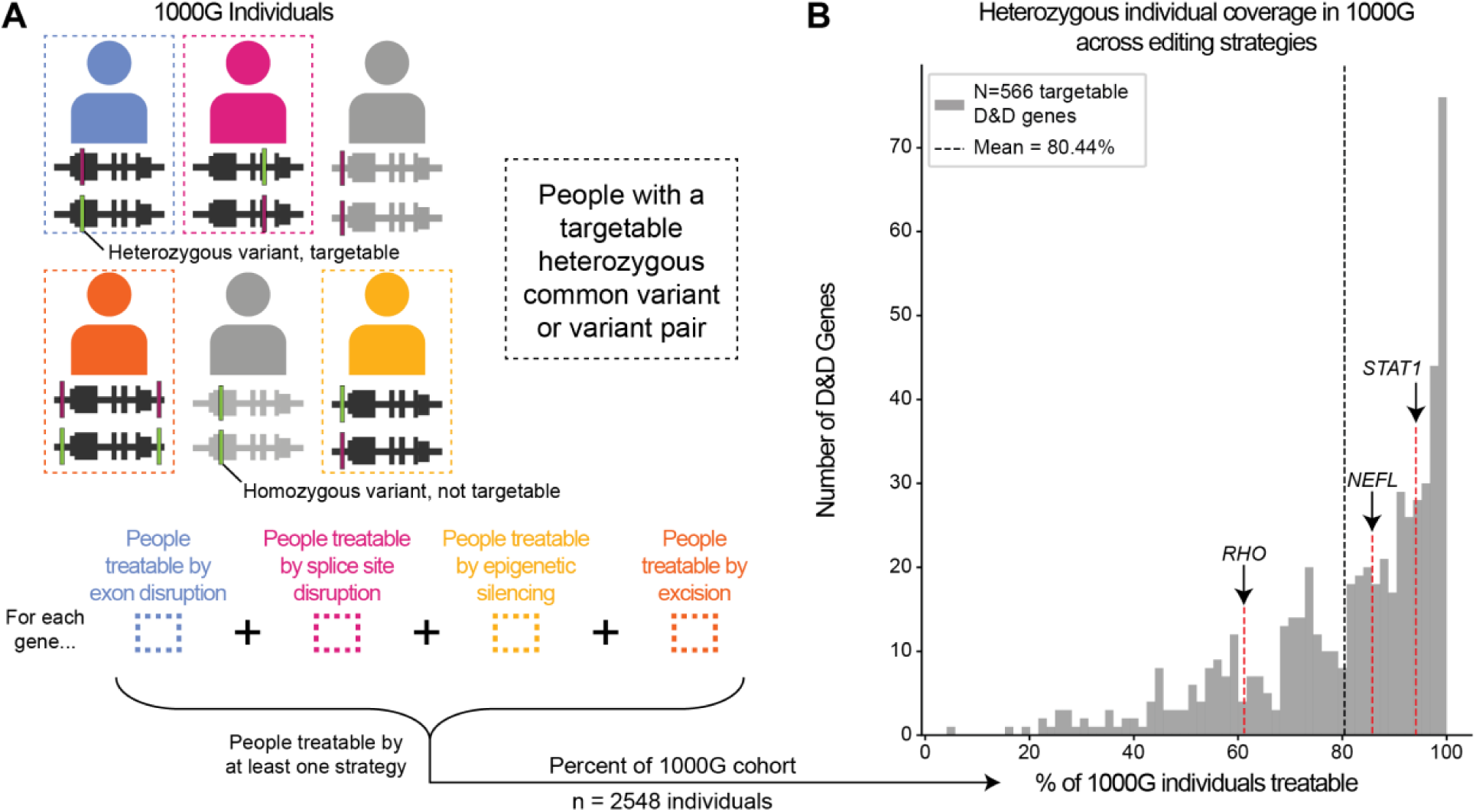
A majority of patients could benefit from targeting common heterozygous variants. (A) Schematic of strategy for identifying individuals who would be treatable based on the presence of heterozygous common variants or variant pairs for each editing strategy around each D&D gene (top). For each gene, we counted the number of people treatable by at least one editing strategy (bottom) to get the percentage of individuals in the 1000 Genomes cohort that would be targetable by our mutation-agnostic allele-specific approach. (B) Percentage of the 1000 Genomes cohort that could have a specific allele targeted by at least one editing strategy for each targetable D&D gene. Across targetable D&D genes, the majority (>80%) of individuals were targetable on average, and the most common scenario across genes was >95% of people targetable. 1000G = 1000 Genomes.

More than 80% of individuals on average had targetable heterozygous variants across D&D genes (Figure 4B). Only 45 D&D genes were targetable in less than 50% of individuals, which could be a suitable threshold for disease interest groups considering therapeutic gene candidates. Breaking down by CRISPR strategy, excisions could treat a median of 82% of people per gene at the high end, and splice site disruption could treat a median of 41% of people per gene at the low end (Supplementary Figure 4B). Thus, assuming that D&D alleles occur randomly across genetic backgrounds, the probability of being able to treat someone by targeting editing to common variants is very high (>80%), illustrating the therapeutic potential of our approach.

### Mutation-agnostic editing of D&D alleles increases per-therapy patient benefit

Given the time and financial barriers to regulatory approval for bespoke therapies targeted to each disease mutation, finding the smallest number of therapies that can target the largest number of patients with disease mutations is crucial for clinical translation. In our approach, different gRNAs are necessary to target common variant alleles. To prioritize common variants for allele-specific targeting by each strategy, we developed a greedy algorithm to select gRNAs matched to common alleles based on their ability to specifically target the largest number of haplotypes in the 1000 Genomes cohort. Subsequent gRNA-allele targeting pairs were selected by choosing those that captured the most haplotypes not targeted by previously selected gRNAs (see Methods). Since humans carry two copies of each chromosome (haplotypes), we considered each separately, to account for the disease mutation possibly occurring on either haplotype. We also filtered targetable common alleles by their proximity to a nearby PAM sequence prior to prioritization to create a pool of selected gRNAs that are experimentally viable. Given that the excision approach requires 2 gRNAs for each edit, we considered each pair of gRNAs as a single therapy for this strategy. We used EXCAVATE-HT (see Methods and Supplementary Figure 6) to confirm the presence of valid PAM sequences and generate gRNA sequences for each common allele. We focused on the CRISPR/SpCas9 system (NGG PAMs), but additional targets could be gained from using other systems. We make all relevant common variants, regardless of CRISPR/SpCas9 targetability, publicly available for viewing through UCSC Genome Browser maps (see Data Availability and Supplementary Figure 7).

To begin with a single-gene example, we describe results for the exon disruption strategy targeting *MYH7*, a myosin heavy chain protein with 149 D&D alleles documented in ClinVar that cause cardiomyopathies. We identified four targetable common heterozygous variants in the exons of the gene for this editing strategy (Figure 5A). The most common targetable allele is G at chr14:23433544 (rs2069540). In the 1000 Genomes population, 24% of the cohort (1,220 haplotypes out of 5,096 haplotypes) would be targetable with a single gRNA therapy. Assuming equal frequency of pathogenic disease mutations, under a mutation-specific editing approach, a single therapy targeting one mutation would only be able to treat 1/149^th^ of the population, or ∼1.3% of haplotypes (Figure 5A). In comparison, the 24% of haplotypes captured by mutation-agnostic editing indicates nearly 18 times as many patients treated, demonstrating that patient benefit could be dramatically improved with this approach.

**Figure 5.**
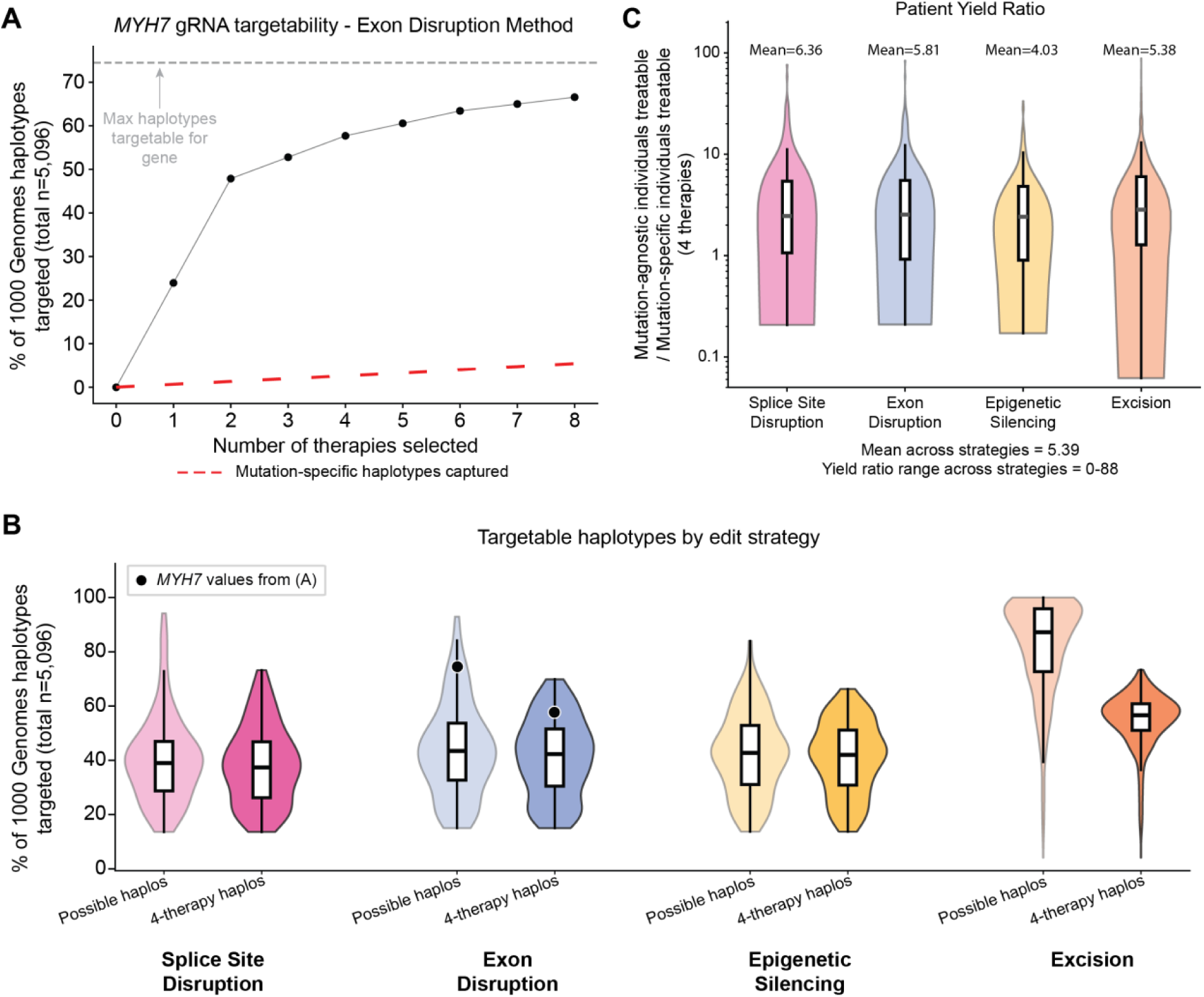
Four therapies would be sufficient to target most haplotypes of heterozygous individuals for many D&D genes, yielding greater patient benefit. (A) Example of the difference in individual haplotypes targetable by disease mutation-specific vs. mutation-agnostic editing strategies for exon disruption of *MYH7*. Nearly 25% of the 1000 Genomes haplotypes are targetable with the first common-variant-targeting therapy. All 8 exon-disruption therapies can cumulatively target ∼67% of cohort haplotypes, which is nearly all of the possibly targetable haplotypes belonging to heterozygous individuals (grey dashed line). The red dashed line represents the number of haplotypes targetable by mutation-specific editing. (B) Percent of total haplotypes in 1000 Genomes that are possibly targetable (denoted as “Possible haplos”) versus that are targetable by the top 4 therapies, denoted as “4-therapy haplos”. The possibly targetable haplotypes are less than 100% for most genes and strategies because some individuals lack targetable heterozygous common variants. For splice site disruption, exon disruption, and epigenetic silencing, the top 4 therapies could target nearly all possible haplotypes, with roughly a ∼2% difference in means between possible haplos and 4-therapy haplos. Excisions could target roughly 55% of the total haplotypes. (C) Yield ratio per gene of the number of treatable people with the top 4 mutation-agnostic therapies compared to 4 mutation-specific therapies. Up to 88 times as many people, and ∼5 times as many people on average across genes, are treatable with 4 therapies compared to mutation-specific gene editing approaches. Only data from genes with more than 4 pathogenic mutations are shown. Note that ratios equal to 0 are not pictured but are factored into the mean values shown.

Across D&D Group 1 genes, only four unique therapies were necessary per gene to treat a large proportion of possible individuals. For exon disruption, epigenetic silencing, and splice site disruption, the top four therapies could target a mean of >38% of haplotypes across D&D genes (Figure 5B), only missing ∼2% of all possible targetable haplotypes (Figure 5B). The excision approach could target a mean of 55% of haplotypes with four therapies (Figure 5B). The targetable fractions were similar for the D&D Group 2 set (Supplementary Figure 5), indicating large proportions of the population would be treatable with a small number of therapies. To directly quantify the patient benefit from the mutation-agnostic approach, we computed the ratio of the number of individuals treatable by the top four prioritized mutation-agnostic therapies relative to the number of patients treatable by four mutation-specific therapies (Figure 5C). A ratio above 1 represents greater patient yield for mutation-agnostic approaches. For example, *MYH7* had an average ratio of 16.7 across editing strategies, indicating nearly 17 times more treatable patients than mutation-specific editing for the same number of therapies. Not every D&D gene has a large number of pathogenic mutations and, therefore, not every gene could gain benefit from mutation-agnostic editing. For example, *SFTPA1*, a surfactant protein in which mutations cause interstitial lung disease, has only 4 dominant mutations, so there is limited benefit of a mutation-agnostic approach with this gene; its average ratio was 0.5. However, for targetable D&D genes with >4 dominant mutations, 5.4 times as many patients would be treatable on average with mutation-agnostic therapies compared to mutation-specific therapies, and >80 times as many patients were treatable for some genes (Figure 5C). These results are likely an underestimate given pathogenic mutations in many genes are still being classified and many remain to be discovered. Thus, mutation-agnostic targeting represents a fundamental improvement over disease mutation-specific approaches, highlighting the large patient benefit that could be gained.

## Discussion

Here we demonstrate that allele-specific editing of D&D genes has the potential to treat hundreds of dominant diseases. Our approach is highly scalable to large proportions of the population, decreasing the burden of regulatory barriers while benefitting a greater number of patients. Moreover, targeting variants common across human populations ensures that genome editing strategies will benefit people of diverse genetic backgrounds.

Our approach is enabled by several conceptual and technical insights. First, we define dominant and dispensable (D&D) genes, a novel clinically relevant class of disease genes, and we identify hundreds of diseases where inactivation of a DN/GOF allele is likely sufficient to restore health. Second, we show that targeting patterns of heterozygous common human genetic variation allows efficient and broadly applicable allele-specific gene editing with only a few gRNA therapies. Third, focusing on common variation for dispensable allele disruption enables the use of diverse editing strategies, making common variant targeting a versatile approach that will adapt and scale with new strategies. While we focus on four main editing strategies, other methods for allele-specific silencing could be integrated into our framework. To support development and adoption of allele-specific therapeutic strategies at scale, we provide genome-wide maps of targetable common variants in and near D&D genes. These maps enable researchers to rapidly evaluate the potential for allele-specific editing of disease genes of interest and promote gene editing therapies for dominant conditions.

Nevertheless, there are limitations to our current work. First, methods for comprehensively determining haplosufficiency are in early stages of development. We relied on large datasets of genetic variation, functional knockouts, and clinical evidence to construct the initial set of D&D genes; however, the annotations we relied on for this are incomplete given current limited understanding of gene and variant function across tissues. Thus, the D&D gene sets will evolve as new annotations become available. Given long-standing interest in haplo*in*sufficiency in clinical genetics, haplosufficiency has much less commonly been directly tested and annotated for dominant genes. As a result, we used imperfect proxies for haplosufficiency, such as the lack of evolutionary constraint against heterozygous loss of function^15^, to refine our gene classifications. Current annotations also fail to capture the dramatic context dependency^30^ and tissue-specificity^31^ that dosage sensitive genes can exhibit. As large-scale tissue-specific gene inactivation methods emerge, it may become possible to accurately determine the physiological contexts in which a gene does or does not exhibit dosage sensitivity, leading to more accurate determination of the potential for tissue-specific therapeutic targeting.

Second, we have incomplete knowledge of rare dominant disease-causing mutations. As a result, the number of dominant pathogenic mutations we identify in D&D genes (Figure 2A) is an underestimate that will increase with advances in genome sequencing and clinical genetics. In addition, we acknowledge that in specific cases, mutation-specific targeting^6,33^ of D&D genes is a viable option that should be considered when there are strong founder effects^32^ or when patients lack heterozygous variants at the gene locus.

Clinical genome sequencing paired with strategies for robust allele-specific gene inactivation provides an unprecedented opportunity to combat diseases caused by D&D alleles. Substantial work lies ahead to achieve this promise. First, refining the definition and understanding of D&D genes will be essential for evaluating the feasibility of therapeutic strategies for specific diseases. Haplosufficiency can vary across biological contexts, and a deeper understanding of the relationship between gene dosage sensitivity and function is needed. Second, integrating the expanding set of powerful genome editing methods into our framework will further increase the potential to silence dominant alleles. For example, base editing has the potential to introduce premature stop codons^34^ in addition to disrupting splice sites, and there is also potential to disrupt non-coding regulatory elements in *cis* with disease alleles^35–37^. Finally, new software is needed to facilitate prioritizing common heterozygous variants and designing the therapies that target them^34^.

Nonetheless, the recognition of D&D genes is already providing a powerful new way to identify and approach potential high-impact therapeutic applications of allele-specific editing^11^. Further advances in clinical genetics and genome editing are likely to increase the number of D&D genes, increase the number of characterized disease mutations per gene, and increase the flexibility of genome editing tools. As a result, the opportunities for allele-specific editing to benefit patients with severe genetic disease will only increase, bringing us closer to delivering gene therapies targeted to the most disease-relevant cellular contexts and ultimately maximizing therapeutic benefit for patients.

## Data Availability

Gene sets and targetability information is included in Supplementary Table 2. Code used to curate common variants and run the greedy algorithm can be found at https://github.com/gramey02/dnd_project, and browser track information can be found at https://github.com/gramey02/DnD_TrackHubs_Public/. Data from ClinGen and ClinVar is publicly available and can be found at https://clinicalgenome.org/ and https://www.ncbi.nlm.nih.gov/clinvar/, respectively. HPO data is also publicly available and can be found at https://hpo.jax.org/data/ontology and https://hpo.jax.org/data/annotations. We curated OMIM data using the OMIM API (https://www.omim.org/help/api) which can be accessed upon request. Data for the 1000 Genomes Project phase 3 release used in this study is also publicly available and can be found at https://www.internationalgenome.org/data-portal/data-collection/phase-3. We used hg38 reference genome alignments.

## ACKNOWLEDGEMENTS

We would like to thank all members of J.A.C.’s and B.R.C’s labs for helpful feedback on this work. G.D.R. was supported by NIH grant F31AG090013-01 and the UCSF Discovery Fellows. J.A.C. was supported by NIH R35GM127087. Q.T.C. and B.L.M. received support from CIRM EDUC4-12766. L.M.J. and B.R.C. received funding from the Charcot-Marie-Tooth Association and NIH R01-NS119678. B.R.C. received support from NIH R01-AG072052 and CIRM INFR6.2-15527. J.A.C. and B.R.C. received support from CIRM DISC0-17363. B.R.C. acknowledges generous support through gifts from the Roddenberry Foundation and Pauline and Thomas Tusher. We also thank Francoise Chanut for valuable editorial and scientific insights. The contents of this publication are solely the responsibility of the authors and do not necessarily represent the official views of NIH, CIRM, or any other agency of the US government or the State of California.

## AUTHOR CONTRIBUTIONS

B.R.C., J.A.C., B.L.M., L.M.J., and G.D.R. conceptualized this research project. G.D.R. investigated and curated data and performed statistical and computational analysis. X.M. contributed to data analysis for enrichment results. Q.T.C. and L.M.J. aided with literature curation. A.G.S. provided EXCAVATE-HT computational workflows and experimental expertise. G.D.R., J.A.C., and B.R.C. wrote the manuscript, and G.D.R. created visualizations. J.A.C. and B.R.C. provided mentorship, review/editing of writing, and funding for the work. Q.T.C., B.L.M., A.S., H.L.W., and L.M.J. provided expertise and manuscript feedback. P.D. and X.M. provided manuscript feedback.

## Supplementary Figures

**Supplementary Figure 1.**
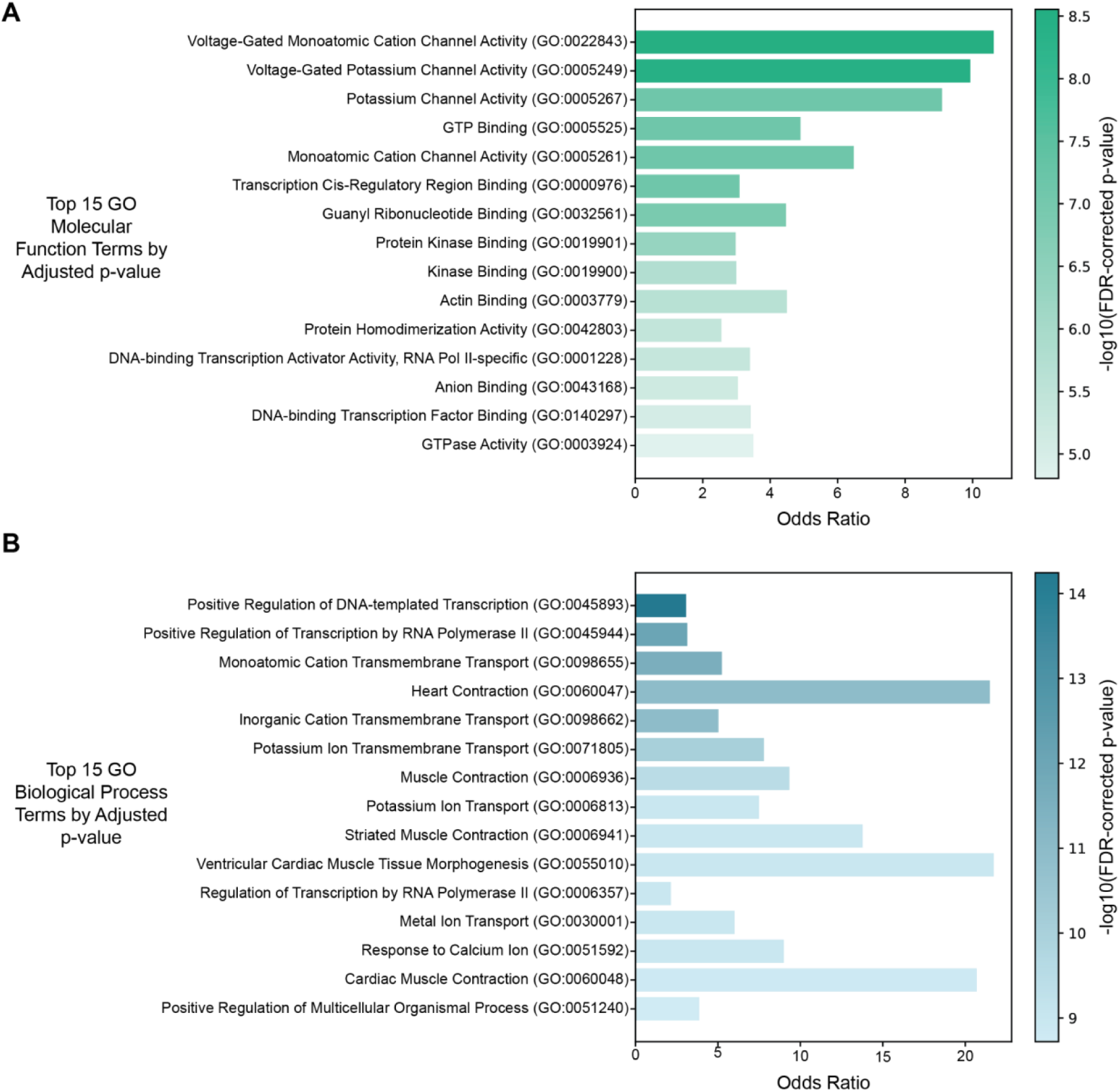
D&D genes exhibit enrichments across many molecular functions and biological processes. (A) Top 15 GO molecular function enrichments among D&D genes. Most prominent terms are related to ion channel activity (consistent with nervous system enrichments in the human phenotype ontology (HPO) catalog), protein and DNA binding, and kinase activity. (B) Top 15 GO biological process enrichments for D&D genes. Most prominent terms are related to ion channel activity, DNA binding, and heart and muscle contraction response, consistent with cardiac system enrichments among HPO terms for the genes. All pictured terms are significant after FDR correction.

**Supplementary Figure 2.**
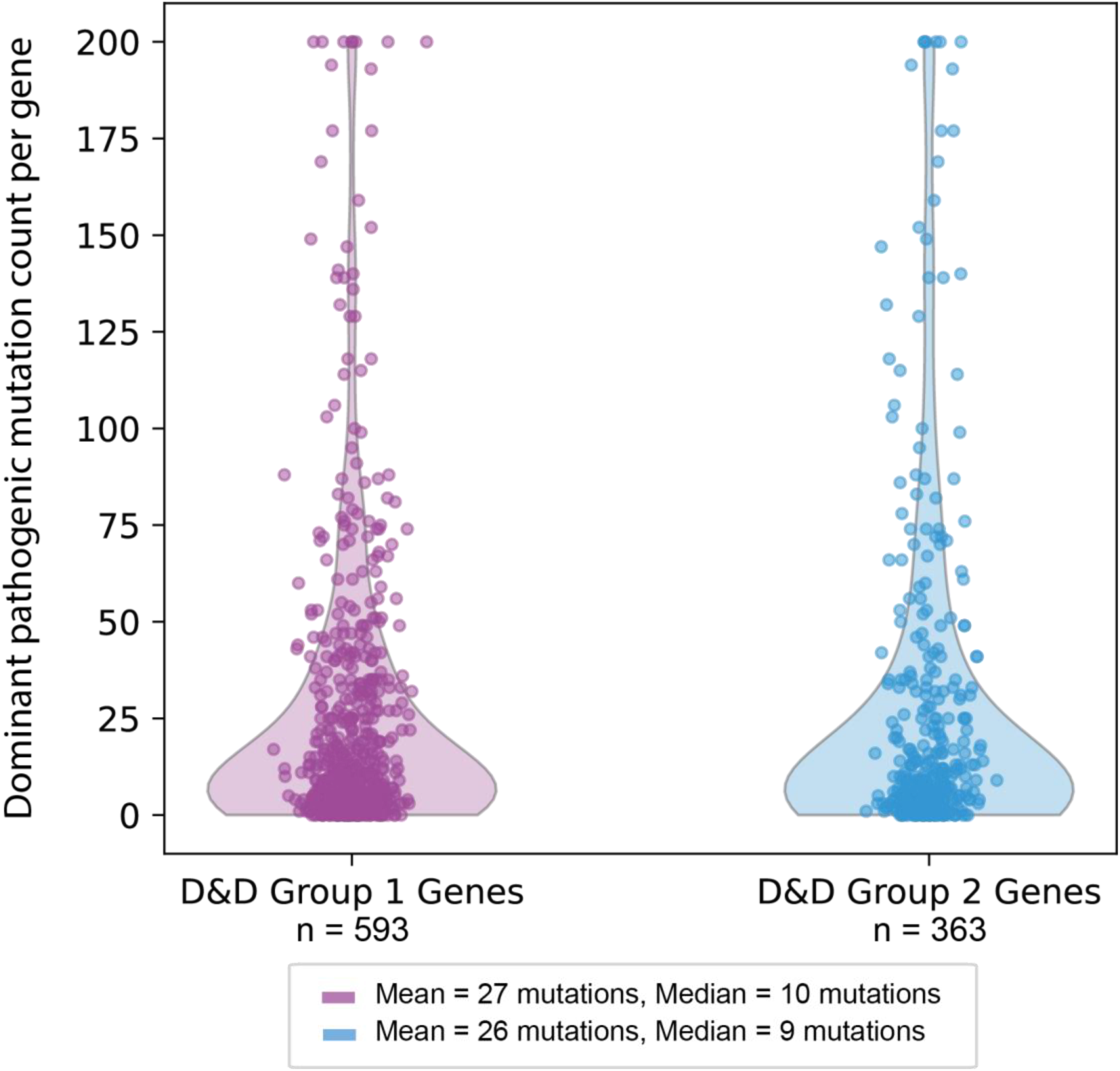
With high pathogenic mutation count, disease mutation-agnostic editing could provide dramatic therapeutic benefit across D&D Group 1 and Group 2 genes. Numbers of dominant pathogenic mutations among D&D Group 1 and Group 2 genes are shown. With high mean and median numbers of disease-causing mutations, mutation-agnostic editing could reach more patients and reduce regulatory burdens for large numbers of therapies. Note that D&D Group 2 genes are a subset of D&D Group 1 genes.

**Supplementary Figure 3.**
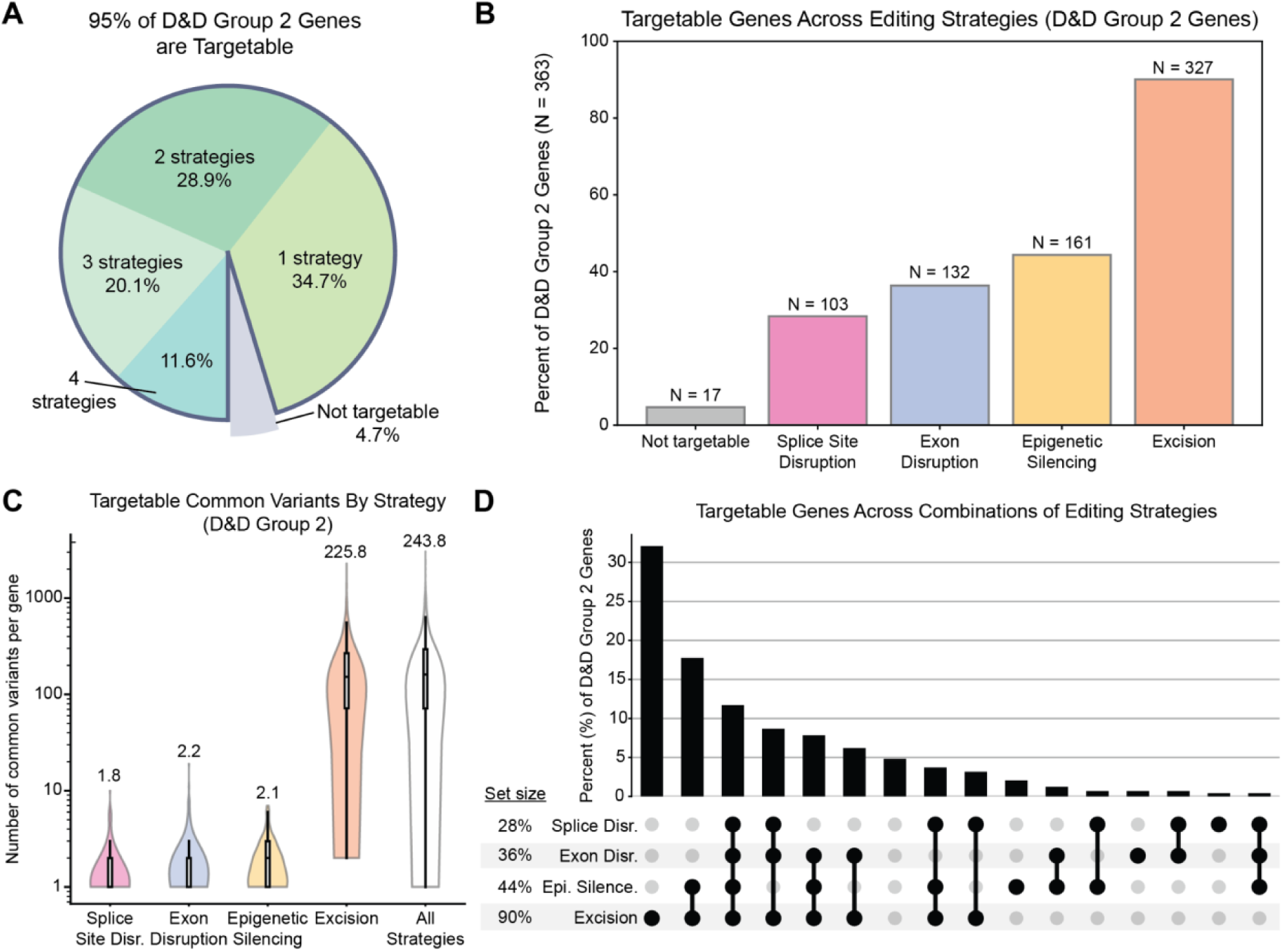
Common-variant-mediated gene editing strategies enable targeting of >95% of D&D Group 2 genes. (A) Over 95% of D&D Group 2 genes were targetable with at least one of the four editing strategies, and over half of the D&D genes (54%) were targetable by more than one strategy. (B) Percentage of D&D Group 2 genes targetable by each editing strategy. Based on common variant patterns, excision could target the most genes (90%), followed by epigenetic silencing (44% of genes), exon disruption (36% of genes), and splice site disruption (27% of genes). (C) Numbers of common variants for each D&D Group 2 gene targetable by each editing strategy, indicating many opportunities to target each gene. Numbers above violins are mean values of each distribution. (D) D&D genes are often targetable by multiple strategies, providing therapeutic versatility.

**Supplementary Figure 4.**
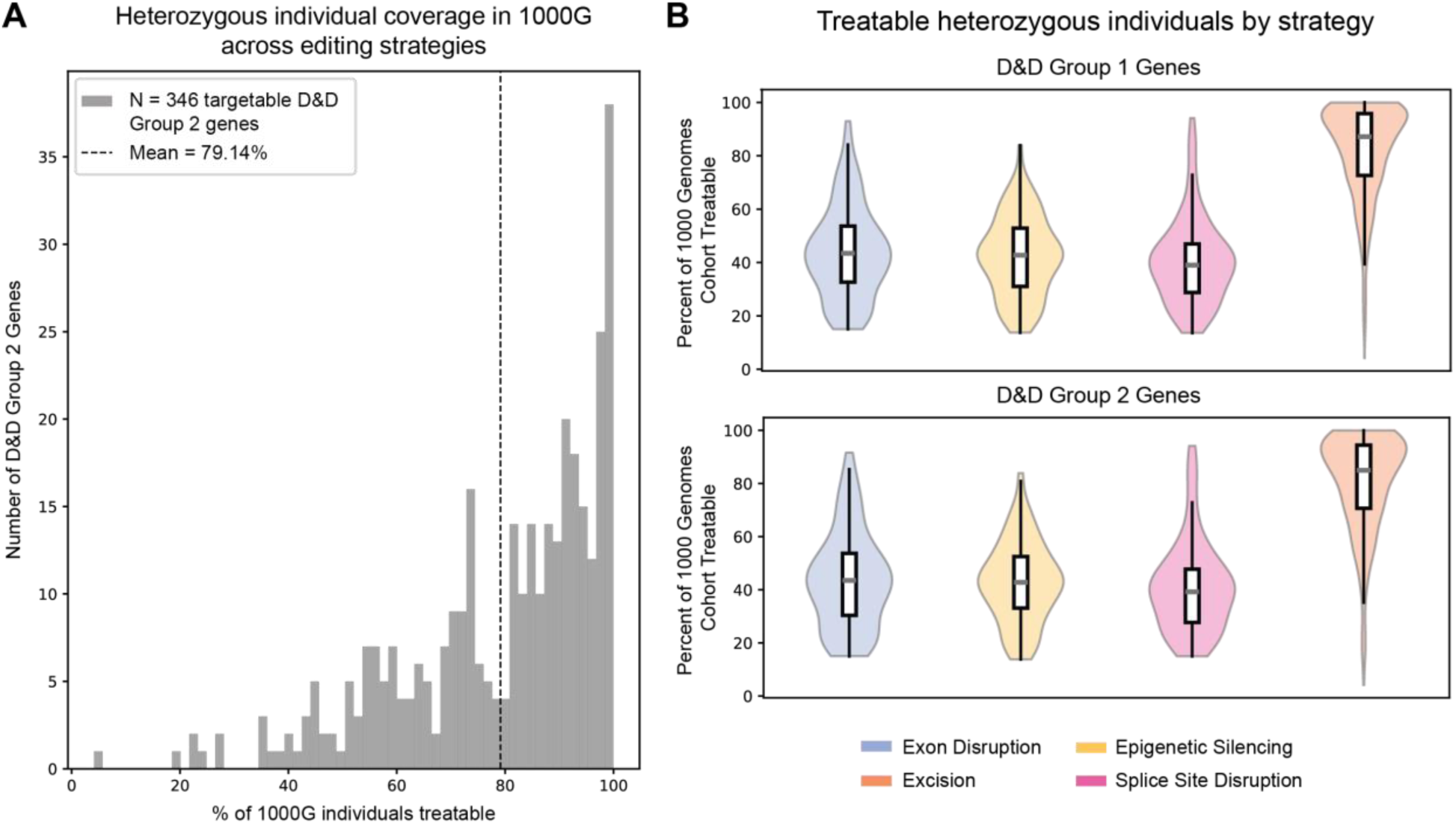
Over 75% of people are therapeutically targetable for D&D Group 2 genes. (A) Percent of total 1000 Genomes cohort that is heterozygous at at least one common variant site for each D&D Group 2 gene, combining unique people across each gene editing strategy. (B) Percent of total 1000 Genomes cohort treatable per gene, broken down by editing strategy for D&D Group 1 Genes (top) and D&D Group 2 Genes (bottom). 1000G = 1000 Genomes.

**Supplementary Figure 5.**
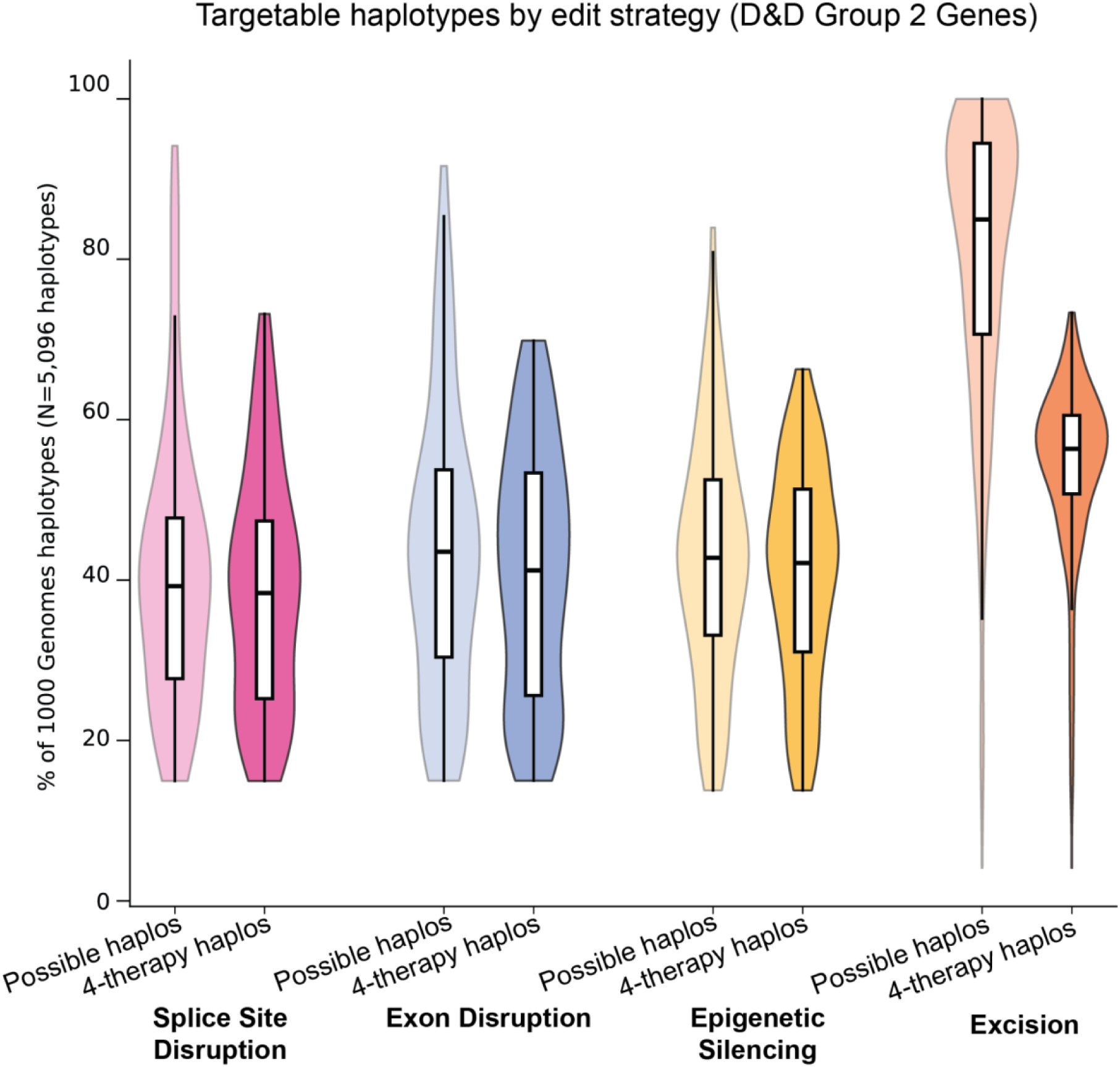
Across a majority of editing strategies, only 4 therapies are required to target a majority of possible haplotypes in the population for D&D Group 2 Genes. Percent of haplotypes out of the total number of haplotypes in the 1000 Genomes cohort that are either targetable at all (by virtue of harboring a heterozygous common variant allele), denoted as “Possible haplos”, or targetable by the first 4 selected therapies, denoted as “4-therapy haplos”. For splice site disruption, exon disruption, and epigenetic silencing, 4 therapies could target nearly all possible haplotypes, with less than a 3% drop off between Possible haplos and 4-therapy haplos. Furthermore, the excision strategy could target 54% of haplotypes on average across genes with only 4 therapies, capturing a large proportion of cohort haplotypes.

**Supplementary Figure 6.**
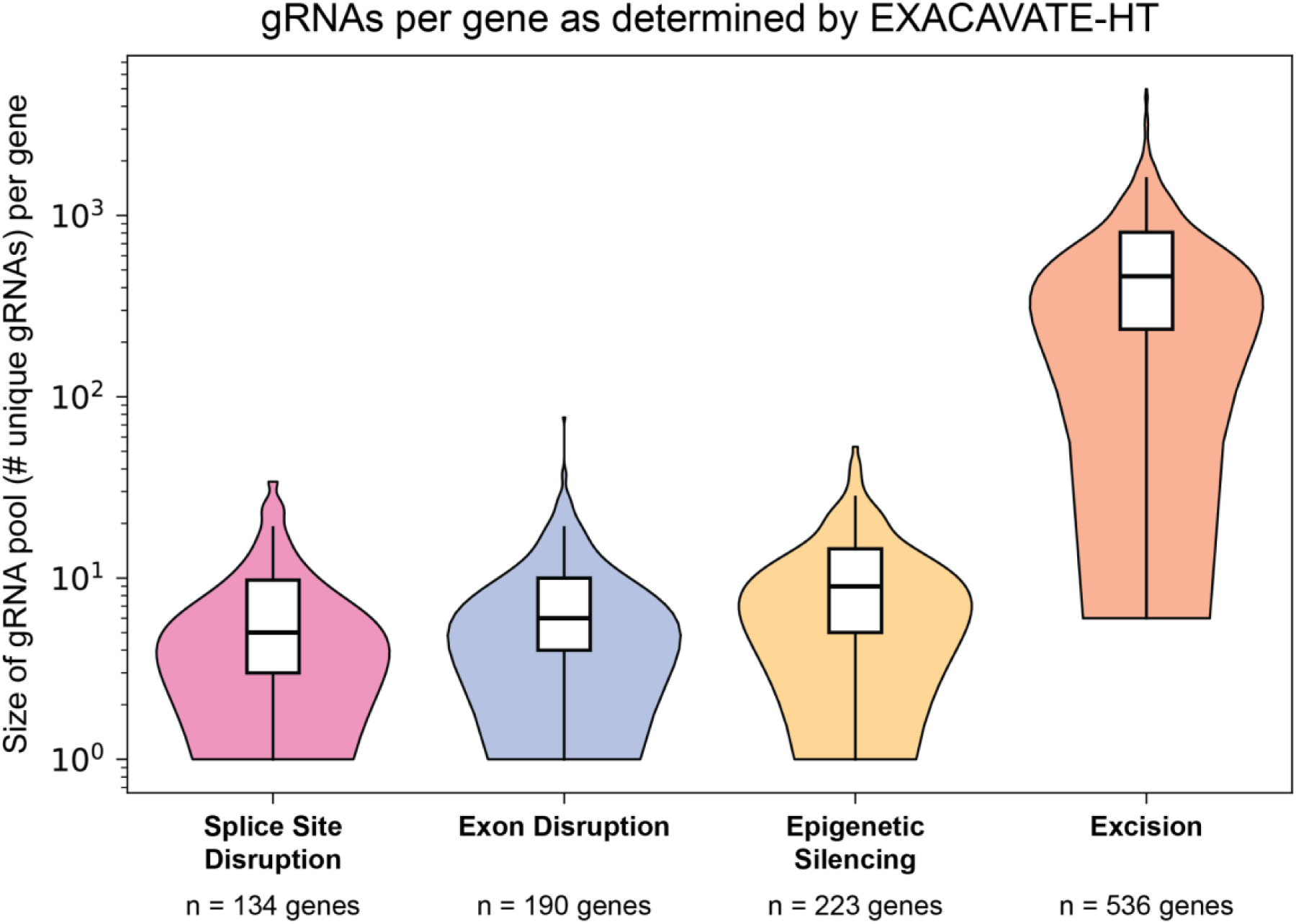
gRNA pools per D&D gene. Number of unique gRNAs available per targetable D&D gene, determined by EXCAVATE-HT. gRNAs were generated by assessing CRISPR/SpCas9 targetability of each common variant and NGG PAM site proximity. Note that numbers of genes targetable by each editing strategy may be lower than in Figure 3C due to some genes not having any common variants targetable by CRISPR/SpCas9.

**Supplementary Figure 7.**
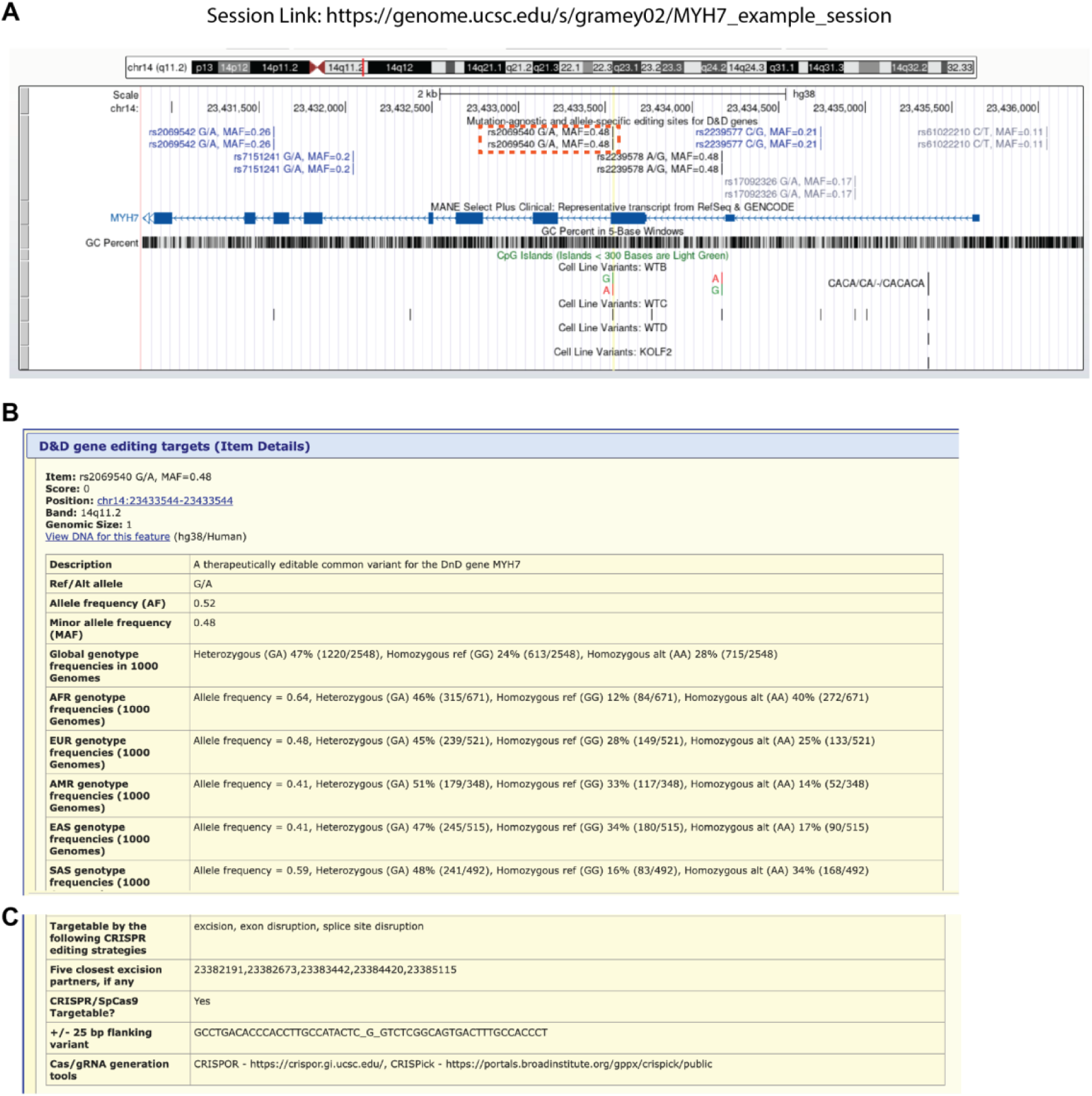
Mutation-agnostic gene editing candidates TrackHub on UCSC Genome Browser. (A) Snapshot of the provided genome browser resource for the gene *MYH7*, with session link that users can click on to be brought to the browser directly (https://genome.ucsc.edu/s/gramey02/ MYH7_example_session). Common variants targetable by mutation-agnostic editing strategies are shown. Note that variants in this window show up twice because the nearby *MYH6* gene is also targetable by excision at some of these sites. Orange dotted box is the common variant highlighted in B & C. (B & C) Snapshots of the clickable inset information provided for each common variant on the track, including subpopulation-specific allele frequencies (B), editing strategy information (C), and flanking sequences (C). Additional tools for gRNA identification are also listed (C).

## Supplementary Tables

**Supplementary Table 1.** HPO and GO Term Enrichments.

**Supplementary Table 2.** D&D gene targetability by edit strategy.

**Supplementary Table 3.** Main figure data.

**Supplementary Table 4.** Manual organ system annotations for missing HPO genes.

## METHODS

### Dominant & dispensable (D&D) Group 1 gene set curation

Gene-disease and gene dosage annotations were last obtained from the ClinGen database (https://clinicalgenome.org/) on November 13^th^, 2025. Genes were selected based on the presence of an autosomal dominant or semidominant annotation with a classification ranging from ‘Moderate’ to ‘Definitive’ in confidence. Genes in the ClinGen database can have more than one mode of disease inheritance linked to them, and we excluded genes with both an autosomal dominant and X-linked annotation while keeping genes with both an autosomal dominant and autosomal recessive annotation. This is to reflect the fact that many genes that exhibit dominant negative- and gain-of-function-based phenotypes can also manifest a recessive form of the same phenotype in the face of homozygous loss-of-function mutations. We included genes associated with “semidominant” conditions because, like full dominance, semidominance can operate via a DN/GOF mechanism^38,39^. We additionally filtered out genes from the D&D set based on the presence of ‘Sufficient Evidence’ or ‘Emerging Evidence’ of haploinsufficiency, keeping genes that had either ‘No evidence of dosage sensitivity’, little evidence/inconclusive haploinsufficiency annotations, or a missing dosage annotation. This resulted in 598 curated genes for the D&D set, 5 of which were filtered out upon transcript filtering (see next section).

## Transcript filtering

We obtained transcript and exon coordinate information on each D&D gene from ENSEMBL version 115. We kept transcripts with the following biotypes annotated in ENSEMBL, as each could viably cause a DN/GOF-linked condition: protein_coding, nonsense_mediated_decay, non_stop_decay, lncRNA, and miRNA. 5 of the 598 D&D genes did not have transcripts that fell into any of these biotypes, reducing the D&D set to a total of 593 genes.

Our main analysis was based on the selection of common heterozygous variants that would inactivate a gene copy, meaning that inactivation across multiple transcripts of the gene would have to be achieved. To ensure we were selecting the most biologically-relevant gene transcripts, we filtered transcripts in several ways. We aimed to remove excessively rare transcripts from consideration by determining how much each transcript contributed to a gene’s expression. We used GTEx v10 data (https://gtexportal.org/home/downloads/adult-gtex/bulk_tissue_expression) to calculate the proportion of expression (in transcripts per million, TPM) each transcript contributed to each tissue the gene was expressed in, excluding cultured cell tissues (’Cells - EBV-transformed lymphocytes’ and ’Cells - Cultured fibroblasts’). Next, we calculated the mean expression proportion of the transcript across the tissues the gene was expressed in. If this mean expression value was less than 5% (i.e., if the transcript contributed on average less than 5% of the total TPM for the gene across tissues), it was removed from consideration.

## shet thresholding and D&D Group 2 gene set curation

We thresholded genes in our D&D set based on the shet metric which has previously been shown to distinguish genes that are under constraint and intolerant to loss-of-function mutations based on a population genetics model and observed genetic variation patterns in the population. Higher values of shet mean a gene is more likely to be under strong constraint and more likely to be haploinsufficient, while a lower shet indicates greater loss-of-function tolerance, and a higher likelihood of being haplosufficient. The metric also accounts for lower statistical power in shorter genes, which we had many of in our gene sets (e.g., *NEFL*).

An shet threshold that distinguishes haplosufficient from haploinsufficient genes has yet to be reported in the literature. We picked a threshold of 0.1 which has been used to delineate genes with a high likelihood of embryonic lethality when harboring a loss-of-function mutation in previous work^15^. We extracted the estimated shet values for each of the 593 D&D genes which are made publicly available (https://zenodo.org/records/10403680), and used it to filter our D&D set to down to 363 genes that passed the filter, which constituted our D&D Group 2 genes.

## CRISPR editing strategies

We analyzed four gene editing strategies for common variant-mediated allele-specific editing: exon disruption, splice site disruption, epigenetic silencing, and excision. Identifying genetic variants targetable by each editing strategy relied on the presence of variants in particular genomic regions (e.g., coding regions, splice donor/acceptor sites, etc.). To ensure any gene editing construct targeted to a variant would have the same effect across all transcripts considered, we only searched for variants in what we deemed “ubiquitous regions” across transcripts. To illustrate the concept with an example, for the exon disruption strategy, variants in exons are of interest, as the goal is to disrupt a gene’s coding sequence. Accordingly, we took the intersection of all exon coordinates across viable transcripts for each gene (see Transcript Filtering), and we searched for common variants in those ubiquitous regions to get the targetable common variants for that gene for the exon disruption strategy. We did this for the specific region of interest across CRISPR editing strategies. For the exon disruption strategy specifically, we only considered common variants as viable strategy targets if they induced NMD across all valid transcripts. We determined if a variant would induce NMD by assuming an indel at the variant locus would induce a protein truncating codon and adhering to the NMD induction rules described by Klonowski et al^40^.

For the splice site disruption strategy, we looked for variants in splice acceptor sites and splice donor sites. Specifically, we looked for variants in the 4-21bp region on either side of a splice acceptor boundary, given the ability to use either a C or A base editor (CBE or ABE, respectively) targeted to the splice acceptor sequence of ‘**A**G’ on either the forward DNA strand or ‘T**C**’ on the reverse strand. For splice donor sites, we searched the 3-21bp region downstream of the splice donor boundary, given the selective ability of CBEs and ABEs to target a splice donor site only on one DNA strand (the ‘GT’ donor sequence is only editable by CBEs and ABEs on the complementary strand, at the corresponding ‘**CA**’ bases). We determined these optimal ranges in which common variants should fall for base editing based on previous work^26^.

For the epigenetic silencing strategy, we defined the promoter region as 500bp upstream and 500bp downstream of the transcription start site per the original CRISPR methylation (CRISPRoff) method^28^. We determined the intersecting promoter regions across viable transcripts per gene and calculated the average GC content across transcripts in the promoter regions. We only looked for common variants in the ubiquitous promoter regions if the average GC content was greater than 30%, as this is a threshold at which others have seen successful CRISPR-based upregulation of methylation at the promoter region^29^.

For the final strategy, excision, we searched for pairs of common variants within a +/-50,000bp window surrounding the boundary of each gene. We shortened the window if other protein coding genes were overlapping in order to retain the largest window not overlapping other crucial genomic elements. We obtained gene body coordinate information from GENCODE release 49. For each gene, we considered pairs of common variants for further analysis only if their coordinates fully encompassed at least one exon across all transcripts considered.

## Common variant identification in 1000 Genomes database

We used data from the 1000 Genomes phase 3 release to identify common variants in the human population that could be targetable by each of the four gene editing strategies. We searched for biallelic variants with a minor allele frequency (MAF) ≥ 0.1 to obtain variants at reasonably high frequency in the population and to ensure these variants would target more than a handful of individuals.

## Identification of nearby PAM sequences using EXCAVATE-HT software

For a common variant to truly be targetable, it is necessary to have a PAM sequence nearby to anchor the Cas nuclease. We determined PAM site proximity using the software EXCAVATE-HT (<u>Ex</u>traction of <u>C</u>ommon <u>A</u>llelic <u>Va</u>riants for <u>T</u>argeted <u>E</u>diting in <u>H</u>igh-<u>T</u>hroughput) which determines PAM locations nearby common variants and corresponding gRNA sequences linked to both standard and custom Cas proteins. We computed our results in EXCAVATE-HT using a CRISPR/SpCas9 construct, meaning we searched for NGG PAM sequences in a 10bp seed region around each common variant of interest. Common variants that had a nearby PAM sequence and therefore were CRISPR/SpCas9 targetable were considered for gRNA prioritization analyses. EXCAVATE-HT also has the ability to accommodate other Cas nucleases and additional targets could be gained from using other systems in the future.

## Guide RNA prioritization with a greedy algorithm

We prioritized and assessed how many gRNAs would be required to target heterozygous individuals in the 1000 Genomes database across each gene editing strategy using a greedy algorithm. Because the 1000 Genomes cohort consists of individuals without the dominant diseases of interest for this study, we considered individuals on a haplotype basis (2 haplotypes per individual). To ensure representation of the full range of human variation, we assumed that rare disease alleles occur across human haplotypes proportional to their frequency in the population. For conditions with strong founder effects, the number of gRNAs needed could be lower. The greedy algorithm ranks gRNAs by how many haplotypes they can target, or “capture”, as follows:

1. Phase each individual’s targetable common variants for a D&D gene into their two haplotypes
2. Of the targetable alleles across common variants, find the allele that targets the most haplotypes in the population
3. Remove those haplotypes and the selected allele from consideration
4. Of the remaining haplotypes in the population, find the allele (of the remaining unselected alleles) that targets the most haplotypes
5. Remove those haplotypes and the selected allele from consideration
6. Repeat these iterative steps until no more haplotypes are gained with allele selections or all alleles have been selected

In the above steps we use allele to represent the targetable nucleotide on either haplotype in an individual, but these can be thought of analogously as gRNAs, since each allele requires its own specific gRNA sequence to be targeted.

Prioritization of alleles (gRNAs) for the excision strategy worked in a similar way, but the greedy algorithm began with the selection of a pair of alleles targeting the most haplotypes in the population. From there, upon each iteration, we selected either one additional allele that could be paired with previously selected alleles, or we selected a new allele pair. We selected either one or two new alleles at each iteration by determining the per-allele haplotype gain in each case (i.e., dividing the number of haplotypes gained by selecting a new pair by 2, and comparing that to the number of haplotypes gained by selecting a single new allele).

## Dominant pathogenic disease mutation analysis

We used data from the ClinVar database to assess how many pathogenic mutations in each D&D gene were linked to a dominant disease. Note that we considered both pathogenic and likely pathogenic mutations, but we term these “pathogenic mutations” throughout the text for brevity. We ran structured API queries to search for pathogenic mutations with a dominant annotation using the autosomal dominant mode of inheritance (“moi”) property feature (https://www.ncbi.nlm.nih.gov/clinvar/docs/properties/). We frequently noticed that mutations that were linked to a disease name that either included the word “dominant” in the title or that were known dominant forms of a disease (e.g., “Charcot-Marie-Tooth disease type 2A2”) were lacking the associated autosomal dominant property annotation. So, to gain a more accurate estimation of dominant pathogenic mutation numbers for D&D genes, we extracted the OMIM IDs linked to each pathogenic mutation and mapped them to their mode of inheritance using the OMIM API. If a mutation had more than one OMIM ID linked to it, we considered it a dominant mutation if any one of the OMIM IDs was associated with autosomal dominance. This gave us a better view of the mutational landscape for our gene sets.

## HPO and GO enrichment analyses

We used data from the Human Phenotype Ontology (HPO) Catalog to get the top-level affected organ system terms for each D&D gene. To limit the phenotype terms to only the dominant conditions affected by each gene, we mapped gene OMIM IDs to autosomal dominant disease OMIM IDs using the OMIM API. We then mapped disease OMIM IDs to their lower-level HPO terms, and we mapped these lower-level HPO terms to the high-level organ system category. 49 genes were missing disease OMIM IDs because of missing inheritance annotations in OMIM, so we manually annotated the affected organ systems of these genes based on clinical presentations listed in OMIM and the literature (manual mappings listed in Supplementary Table 4). For the genes where disease OMIM ID to HPO term mapping was available, we only considered HPO terms that reached 80% frequency in patients, i.e., if the fractional frequency was >= 80% or if it was listed as HP:0040281 ("Very frequent (99-80%)") or HP:0040280 ("Obligate (100%)"). We also allowed terms to pass if they had no frequency listed, to ensure lack of annotation did not affect the phenotypic proportions we observed. We computed the overall percentage of D&D genes linked to each top-level term. To determine significant enrichment of specific terms in our gene set, we ran a permutation test. We randomly selected 100 sets of 200 genes at a time from the HPO catalog and computed the percentage of genes a top-level term appeared in out of all genes in each permutation set. We computed the mean of each top-level term’s distribution of percentages across all permutations, and we compared that to the observed percentage for the term in our D&D gene sets to obtain an effect size and p value. We used a Benjamini-Hochberg correction within each D&D group enrichment test (e.g., one for D&D Group 1 p values and one for D&D Group 2 p values) to obtain corrected significance outputs.

For GO enrichments, we used the EnrichR tool (https://maayanlab.cloud/Enrichr/) to generate biological pathway and process enrichments for all D&D genes. Reported results are using the default background set of genes for comparison. Full enrichment results are reported in the Supplementary Figure 1 and Supplementary Table 1.

## Patient Yield Ratios

To get a quantitative sense of the benefit of mutation-agnostic editing over mutation-specific editing, we computed a ratio of the number of patients captured by each approach for the first 4 gRNAs of each editing strategy. We calculated the number of mutation-specific treatable people per gene by dividing the total people in the 1000 Genomes cohort by 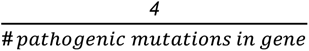. This assumes random distribution of D&D alleles across the population and determines the number of people treatable by 4 therapies. For the mutation-agnostic approach, we considered individuals treatable only if both of their haplotypes were targeted by the first 4 therapies prioritized by our greedy algorithm. We calculated the final ratio by dividing the number of mutation-agnostic people by the number of mutation-specific people.

## UCSC Genome Browser TrackHub

The track hub that readers can use to view all targetable common variants across all D&D genes can be found at https://raw.githubusercontent.com/gramey02/DnD_TrackHubs_Public/refs/heads/main/b ed_files/metadata/All_DnD_genes_hub_file_wCellLines_ng.txt. This link can be loaded into the TrackHub viewer for use. Saved session views to be directly brought to the hub can be accessed at https://genome.ucsc.edu/s/gramey02/MYH7_example_session and https://genome.ucsc.edu/s/gramey02/All_DnDgenes_session. Single gene hubs and bigBed files for viewing targetable common variants can be found at this GitHub repository: https://github.com/gramey02/DnD_TrackHubs_Public/. Data is shown in conjunction with commonly used cell line variants.

## Code and data availability

Gene sets and targetability information is included in Supplementary Table 2. Common variants sets can be visualized in the provided UCSC Genome Browser TrackHub (see previous section). Code used to curate common variants and run the greedy algorithm can be found at https://github.com/gramey02/dnd_project, and browser track information can be found at https://github.com/gramey02/DnD_TrackHubs_Public/. Data from ClinGen and ClinVar is publicly available and can be found at https://clinicalgenome.org/ and https://www.ncbi.nlm.nih.gov/clinvar/, respectively. HPO data is also publicly available and can be found at https://hpo.jax.org/data/ontology and https://hpo.jax.org/data/annotations. We curated OMIM data using the OMIM API (https://www.omim.org/help/api) which can be accessed upon request. Data for the 1000 Genomes Project phase 3 release used in this study is also publicly available and can be found at https://www.internationalgenome.org/data-portal/data-collection/phase-3. We used hg38 reference genome alignments.

